# Alterations in relative perfusion and synaptic patterns in Parkinson’s disease using dynamic SV2A PET

**DOI:** 10.64898/2026.09.28.26364237

**Authors:** Praveen Honhar, Sule Tinaz, Faranak Ebrahimian Sadabad, Jean-Dominique Gallezot, Mika Naganawa, Mark Dias, Yunpeng Ye, Hong Gao, Nabeel Nabulsi, Yiyun Huang, Sophie E Holmes, Robert A Comley, Ansel T Hillmer, Sjoerd J Finnema, David Matuskey, Richard E Carson

## Abstract

**Purpose:** Dynamic [^11^C]UCB-J PET can simultaneously provide information about synaptic density and relative perfusion in Parkinson’s disease (PD). We investigated whether these complementary biomarkers could explain a higher percentage of motor symptom severity variation in PD and inform other PD-specific physiological alterations and symptomatology.

**Methods:** Binding potential (*BP*_ND_) and relative perfusion (*R*_1_) images were derived 30 PD and 30 controls imaged with [^11^C]UCB-J PET. A spatial covariance analysis was used on *R*_1_ to derive PDRP-a known metabolic network alteration in PD. Linear models related PD-motor symptom severity to PDRP expression and nigral synaptic losses. Next, whole-brain cluster analyses identified physiological correlates for PD-tremor severity, a prominent motor symptom that does not correlate with dopaminergic losses. Lastly, inter-individual regional correlation alterations in PD were examined for both *BP*_ND_ and *R*_1_.

**Results:** PDRP was identified from *R*_1_, with higher expression in PD (*p*=1.7×10^-5^, Cohen’s *d*=1.2), and its expression correlated with MDS-UPDRS III (*r*=0.57, *p*=0.0009). Combining PDRP expression scores with synaptic density in the substantia nigra explained 48% of the total variation in MDS-UPDRS Part III, twice as much as previously reported using nigral synaptic density alone. Two significant large clusters (*p*_k_<1×10^-6^) were identified in the occipital-parietal areas where *R*_1_ was negatively correlated with tremor severity (*r* ∼ -0.65, *p*<0.001). Extensive correlation alterations were observed in synaptic density (prominently involving pallidum) and perfusion (prominently involving thalamus).

**Conclusions:** Combining synaptic and perfusion outcomes for [^11^C]UCB-J PET explains more physiological alterations associated with PD.

## Introduction

Parkinson’s disease (PD) is currently the fastest growing neurodegenerative disorder[1] and more *in vivo* imaging biomarkers are needed to expand our understanding of disease pathobiology and testing the efficacy of novel therapeutics in future trials. α-synuclein, whose aggregates are the primary pathology of PD[2], co-localizes pre-synaptically[3] and is involved in SNARE complex assembly[4]. Leading theories about the toxicity of Lewy bodies are based on their interactions with pre-synaptic proteins[5], therefore, pre-synaptic alterations are amongst the earliest changes associated with PD pathology[6].

On the imaging side, the development of the PET radiotracer [^11^C]UCB-J[7] and its variants[8] (including [^18^F]-versions[9, 10]), have made *in vivo* quantification of synaptic density possible[11-14]. These compounds bind to synaptic vesicle glycoprotein 2A (SV2A), a protein expressed on all synaptic vesicles that correlates with other synaptic markers[11]. A few studies have already reported on the emerging applications of this imaging target in measuring nigrostriatal synaptic losses in PD[15-21]. Our previous study with this tracer also reported a significant negative association between nigral synaptic density and motor symptom severity in PD[21]. Along with the measure for synaptic density (binding potential, *BP*_ND_), modeling of full dynamic PET data can also be used to compute a secondary outcome measure that is a surrogate for relative perfusion (*R*_1_). As early time-window PET data is often uncollected to keep scan-times short, *R*_1_ is an underutilized outcome in PD.

This study used complementary synaptic density (*BP*_ND_) and relative perfusion (*R*_1_) measures derived from dynamic [^11^C]UCB-J PET to investigate synaptic and physiological alterations in PD. First, we tested whether the ‘Parkinson’s disease-related motor pattern’ (PDRP) – a widely reproduced abnormal metabolic network that is overexpressed in PD and correlates strongly with motor symptoms[22-25] – could be identified in our cohort with the [^11^C]UCB-J PET outcomes. PDRP is typically derived by application of the scaled subprofile model (SSM) - a principal component analysis (PCA) based spatial-covariance method[26], to metabolic or blood-flow neuroimaging datasets. We further examined whether PDRP expression and nigral synaptic density provide complementary information about motor severity. Second, given that tremor severity in PD is uncorrelated with dopaminergic losses[27], and evidence for the role of brain structures outside the nigrostriatal system in tremor pathophysiology[28], we performed whole-brain cluster analyses to identify brain voxels that correlated with tremor-scores in our cohort. Third, an exploratory analysis was performed to simultaneously identify group-level alterations in regional synaptic (*BP*_ND_) and perfusion (*R*_1_) correlations in PD. The correlations in this analysis represent the across-individual associativity, across subjects, resulting in identification of region-pairs for which the group-level difference in Pearson’s *r* was significantly altered. To our knowledge, none of these analyses have been previously reported with [^11^C]UCB-J PET.

## Methods

### Study Participants

Our study cohort is the same as the one reported in a recent study[21], and is briefly summarized here. Thirty individuals with PD (mean ± SD age = 63±8; 17 women, 13 men, Hoehn-Yahr: 2) and 30 healthy controls (mean ± SD age = 61±9; 17 women, 13 men) were recruited as a part of a prospective PD study at the Yale PET Center (see Table 1 for complete demographic and radiotracer injection details). PD diagnosis was confirmed by the Movement Disorders Society (MDS) criteria[29]. All participants were examined for medical history, physical and neurological examination, and underwent routine blood tests. Exclusion criteria included any history of neurological, neuropsychiatric or addiction-based disorders (other than PD). The study was approved by Yale University Human Investigation Committee and Radiation Safety Committee and all participants signed informed consent forms.

**Table 1:** Clinical characteristics of the cohorts and radiotracer injection details. (Mean ± SD) reported.

|  | Healthy controls | Parkinson's disease | p-value |
| --- | --- | --- | --- |
| N | 30 | 30 | - |
| Age (years) | 61.0 ± 8.8 | 62.8 ± 7.7 | 0.40 |
| Sex (Female:Male) | 17:13 | 17:13 | - |
| MDS-UPDRS III | - | 29.7 ± 8.9 | - |
| MDS-UPDRS Total | - | 48.4 ± 16.8 | - |
| Tremor severity score | - | 5.3 ± 4.5 | - |
| Hoehn & Yahr | - | 2.0 ± 0.0 | - |
| Disease duration (years since symptom onset) | - | 4.6 ± 3.0 | - |
| Injected dose (MBq) | 577 ± 163 | 580 ± 158 | 0.95 |
| Injected mass (ng/kg) | 23 ± 17 | 21 ± 19 | 0.60 |

### Clinical Assessments

Clinical assessment of motor burden of the disease was made through the administration of MDS-Unified Parkinson’s Disease Rating Scale[30] Part III (MDS-UPDRS Part III) in the medication off state. In addition to total motor severity, we also calculated the tremor severity scores which was computed as the sum of items 3.15 to 3.18 measuring the rest tremor amplitude and constancy of the limbs, lips/jaw and postural and kinetic tremor amplitude of the hands. The mean±SD of MDS-UPDRS Part III for our cohort was 30±9 (range: [14-49]), and the same values for the tremor scores were 5.3±4.5 (range: [0-14]). Stebbins et al.[31] validated a criterion based on the following cutoffs for the ratio of mean tremor score to mean postural instability/gait difficulty (PIGD) score (items 3.10 to 3.12 on MDS-UPDRS III) to identify tremor dominant phenotype in PD: ratio ≥ 1.15 (tremor-dominant), 0.9 to 1.15 (indeterminate) and ≤ 0.9 (PIGD dominant). Based on this classification, our cohort was composed of 18 tremor-dominant, 9 PIGD-dominant and 3 indeterminate motor phenotypes of PD.

### Positron Emission Tomography with [^11^C]UCB-J and Magnetic Resonance Imaging

All participants underwent a dynamic PET imaging session for 60 min on the High Resolution Research Tomograph (HRRT) after a one-minute bolus injection (injected activity:579±160 MBq, injected mass:22±18 ng/kg) of [^11^C]UCB-J PET. PET data were reconstructed using the MOLAR algorithm with corrections for attenuation, scattered and random coincidences, detector dead time and head motion[32] (using optical tracking via Polaris Vicra, NDI Systems, Waterloo, Canada). All PET scans were performed in the medication off state (no medications for at least 12h before scans), consistent with clinical assessments.

Participants also underwent a T1-weighted 3T MRI scan with the magnetization-prepared rapid acquisition gradient echo (MPRAGE) sequence with following parameters: echo time (TE): 2.44 ms, repetition time (TR): 2500 ms, inversion time=900 ms, flip angle=9°, voxel size=1 mm^3^ isotropic).

### Image Analysis: PET Quantification

Voxel-wise PET time activity data were fitted to the simplified reference tissue model (SRTM2)[33] with a 2mL volume of the centrum semiovale used as the reference region [34], to generate parametric images of non-displaceable binding potential (*BP*_ND_, measure of synaptic density) and relative delivery parameter (*R*_1_, measure of relative blood-flow).

### Identification of PDRP

PET outcome images (*BP*_ND_ or *R*_1_) from all subjects were first spatially normalized to the Montreal Neurological Institute (MNI) 2mm space and smoothed with a Gaussian filter (Full Width Half Maximum [FWHM]: 5mm). The Scan Analysis and Visualization Processor toolbox, developed at Feinstein Institute for Medical Research and implemented in MATLAB, was used to perform the SSM-PCA analysis[26], consistent with previous reports[23-25, 35, 36]. Briefly, images were masked to remove out-of-brain and low-activity voxels using an optimized value of 30% of maximum voxel value as the lower threshold (see, supplementary file for optimization details), log-transformed and then centered by sequentially removing subject and group means. A PCA analysis was performed on the residual voxel values leading to identification of several principal components (PCs). Out of these, the first few components that cumulatively explained up to 50% of the total variation in the data, and whose expression *z*-scores were significantly different (*p* <0.05) between PD and HC, were retained for further analysis. Then *stepwiseglm* was used in MATLAB v2023b with the default *F*-test criteria to investigate if a linear combination of retained PC scores better separated PD from controls using logistical regression. These steps have been previously recommended for PDRP identification[23].

### Effect of Image Smoothing and an Alternate (Gray Matter) Mask on PDRP Identification

Image smoothing is applied when normalizing PET images to a template space like MNI. As our PET data were collected on a high-resolution tomograph (HRRT), we only smoothed to a FWHM of 5mm to retain finer features. However, most studies identifying PDRP with FDG PET have traditionally used larger smoothing kernels (FWHM 8-10 mm) and image acquisitions on tomographs with lower resolution[23-25, 36, 37]. To compare how our smoothing kernel may impact the findings compared to literature estimates, especially in % variation captured by PCs, we performed a secondary SSM-PCA analysis with a higher level of smoothing (FWHM: 10mm) on *R*_1_ images. Similarly, to investigate whether the masking strategy employed in the main analysis (using an optimized lower threshold of 30% of maximum value) is better than using a gray matter mask, we also performed an SSM-PCA analysis to identify PDRP in voxels identified as gray matter (p_GM_ > 0.3, SPM12 used for segmentation, 5 mm FWHM Gaussian smoothing).

### Combined Effects of Nigral Synaptic Density and PDRP Subject Expression Score on Motor Severity

To test whether the identified PDRP subject expression scores from *R*_1_ images (see Results) provides a complementary explanatory variable for motor severity scores (MDS-UPDRS Part III) compared to the previously established[21] significant negative association between synaptic density of the substantia nigra and the MDS-UPDRS Part III, we implemented the following stepwise general linear model:

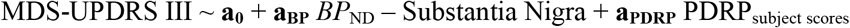

The model started with only the constant term and could add or remove both predictors based on Akaike information criterion (AIC).

### Cluster Analysis for Brain Correlates of Tremor Severity

Using the spatially normalized PET outcome images (*BP*_ND_ or *R*_1_ with FWHM: 5mm Gaussian smoothing), whole-brain voxel-wise correlations (across-subjects) of tremor severity scores with PET outcomes were performed resulting in 3D maps of Pearson’s ***r*** and accompanying *p*-values. A threshold corresponding to *p*_uncorrected_ < 0.01 (corresponding to | ***r*** | > 0.46) was applied to Pearson’s ***r*** image. Next, a cluster analysis was performed on the thresholded image using FSL’s *cluster* algorithm, and only clusters with considerable spatial extent were considered significant (k > 100 voxels, *p*_k_ < 0.0001; *p*_k_: Gaussian random field theory estimate of the probability of cluster size k due to random chance after accounting for spatial resolution of the raw Pearson’s ***r*** image using FSL’s *smoothest* function). Scatter plots of mean PET outcome values within each significant cluster and tremor scores were created.

### Alterations of regional synaptic (BP_ND_) and perfusion (R_1_) correlations in PD

Regional mean values of PET outcomes measures were calculated for all subjects for 22 brain regions of interest (ROIs)– cerebellum, brainstem, substantia nigra (SN), red nucleus, locus coeruleus (LC), raphe nucleus, pallidum, caudate, putamen, ventral striatum, subthalamic nucleus (STN), thalamus, amygdala, parahippocampal gyrus, posterior cingulate cortex (PCC), anterior cingulate cortex (ACC), precentral, supplementary motor area (SMA), postcentral, olfactory cortex, orbitofrontal cortex and ventromedial prefrontal cortex as described previously[21].

Regional Pearson’s correlation matrices (22×22 symmetric matrix, correlations calculated across-subjects) were computed for binding potential (*r*_BP_) and relative perfusion (*r*_R1_) separately in the PD and control cohorts. These matrices define the synaptic and perfusion (*R*_1_) correlations at the group level in both populations. Next, the alterations in pairwise-regional correlations between the two groups was calculated as, *d*_X_ = *r*_X, PD_ – *r*_X, HC_, where X=*BP* or *R*_1_. For inference testing, a permutation analysis (10,000 repeats) was conducted by dividing the 60 subjects randomly in two equal groups and computing the difference in correlations for each region-pair under the null hypothesis of no group difference. For each region-pair, *p*-value for the difference in correlation was defined as the fraction of permutations that showed a more extreme difference in correlation by chance, compared to the observed difference in correlation between the PD and control groups. Per the exploratory nature of this analysis, no adjustment for multiple comparisons was performed (results reported at two-tailed *p* < 0.05 uncorrected level).

## Results

### Identification of PDRP Network

For *R*_1_, the SSM-PCA method identified three PCs (PC1: 7.4%, PC4: 4.2% and PC8: 2.6% variation explained) that showed a statistically significant difference (*p* <0.05) between PD and controls. However, *stepwiseglm* only retained PC1 in the logistical regression and therefore, PC1 was the sole candidate for PDRP (Figure 1a). The subject expression *z*-scores of PC1 were significantly different between PD and HC [mean(SD) PD group: 0.52(0.97), mean(SD) HC group: -0.52(0.73), *p* (2-sample, 2-tailed *t*-test) = 1.7×10^-5^, Cohen’s *d*=1.2, Figure 1b]. The loadings for PC1 were positive (higher) in the voxels within the cerebellum, pallido-thalamic and pontine regions and negative (lower) in the voxels corresponding to parieto-occipital and inferior parietal areas, consistent with the topography of PDRP. Further, within the PD cohort, subject expression *z*-scores for this network were significantly correlated with motor severity (MDS-UPDRS Part III) scores (Pearson’s *r* = 0.57, *p* = 0.0009, Figure 1c). For *BP*_ND_, the SSM-PCA analysis did not reveal any PC that could significantly separate PD from controls.

**Fig. 1.**
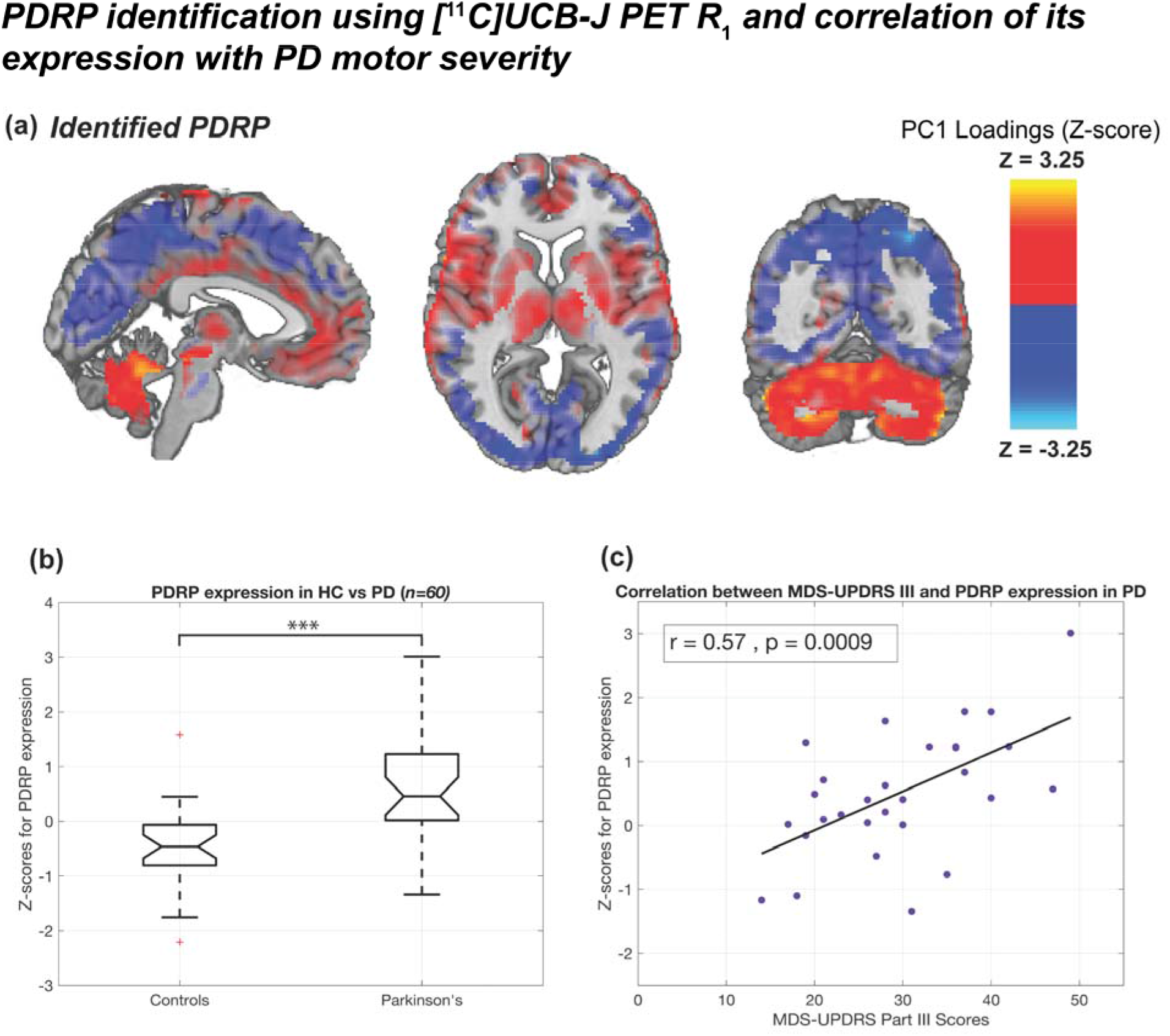
(a) PDRP identified using [^11^C]UCB-J PET *R*_1_ values. The known topography of the network with positive loadings in the cerebellum, pallidothalamic and pontine regions and negative loadings in the occipital-parietal regions, with higher expression in PD, was observed. (b) Boxplots showing significantly higher PDRP expression in PD, compared to controls (*** *p* < 0.0001). (c) MDS-UPDRS Part III scores were significantly associated with *z*-scores for PDRP expression in PD subjects.

### Effect of Image Smoothing and an Alternate (Gray Matter) Mask on PDRP Identification

Higher smoothing of *R*_1_ images (FWHM: 10mm) increased the % variation explained for the PDRP network to 11.8%. The pattern’s topography was similar to the PDRP derived from 5mm FWHM *R*_1_ images (not shown), but while the separation of PD and controls was still robust (*p* < 0.001), the correlation between PDRP expression score and MDS-UPDRS Part III (Pearson’s *r* = 0.50, *p*=0.005) was weaker for 10 mm FWHM Gaussian smoothing.

Using an SPM12-derived gray matter mask for MNI 2mm template space independently increased the % variation explained for the derived PDRP network to 10.5%. However, while the PDRP derived based strictly on gray matter segmented voxels still distinguished PD from controls (*p* < 0.001), PDRP expression score derived through this analysis was uncorrelated with MDS-UPDRS Part III scores (Pearson’s *r* = 0.11, *p* = 0.56). This is likely due to erosion of voxels in key subcortical areas when using a gray matter mask, prominently within the thalamus and globus pallidum.

### Combined Effects of Nigral Synaptic Density and PDRP Subject Expression Score on Motor Severity

The best model for MDS-UPDRS Part III included *both* predictors, *BP*_ND_-Substantia Nigra and PDRP_subject scores_, with the following estimated values (s.e, *p*-value) for the parameters: **a**_**0**_: 36.3 (3.6, *p*<1×10^-9^), **a**_**BP**_: -12.0 (4.2, *p*=0.008), **a**_**PDRP**_: 0.47 (0.11, *p*=0.0002). This model had an AIC value of 202.9, while a model with just the intercept and PDRP_subject scores_ had an AIC of 208.7. The model with both predictors had an R^2^ of 0.48 (Figure 2a), while our previous [^11^C]UCB-J PET study[21] reported an R^2^ of 0.23 with *BP*_ND_-Substantia Nigra. Further, *BP*_ND_-Substantia Nigra and PDRP_subject scores_ were uncorrelated predictors (*p*=0.80, Figure 2b).

**Fig. 2.**
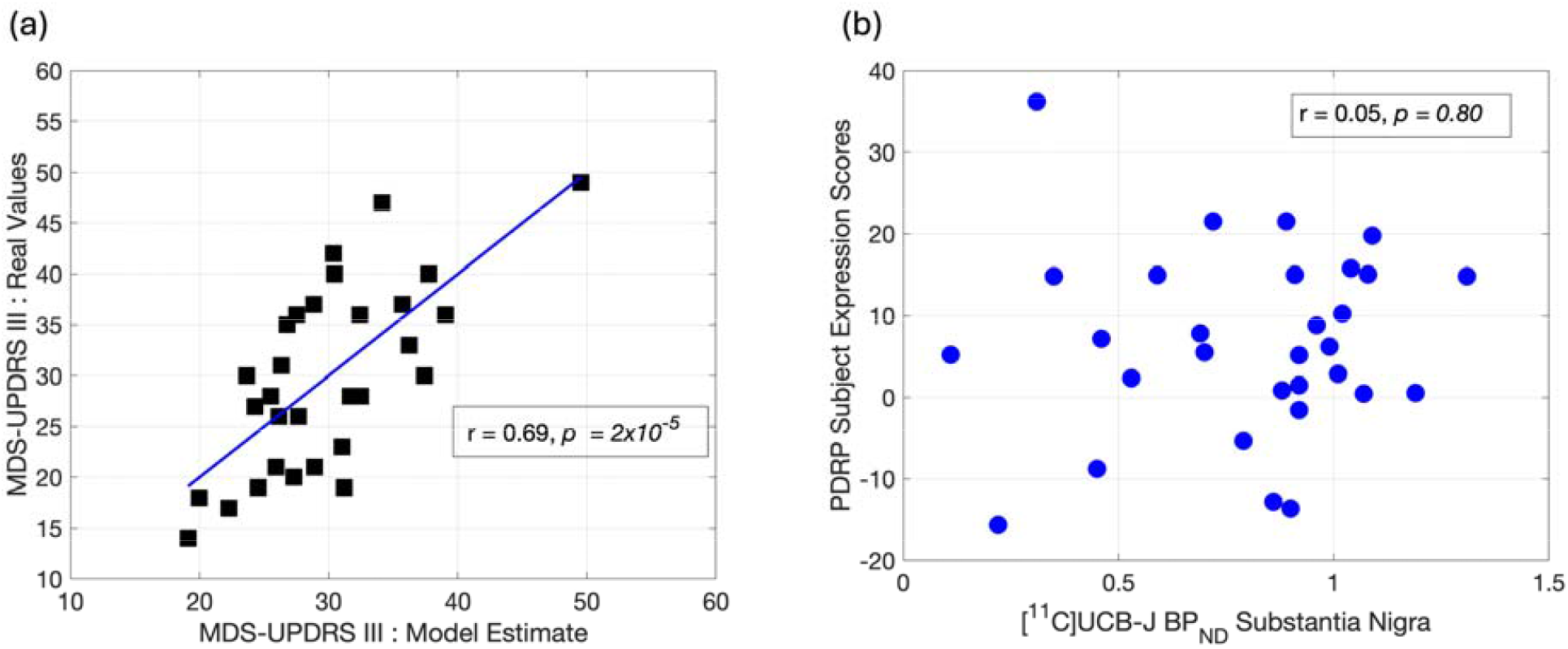
(a) Linear model for MDS-UPDRS III scores that combines synaptic density in the substantia nigra with PDRP subject expression scores together as predictors explain more variability in real data. (b) Scatter plots of binding potential in the substantia nigra and PDRP subject expression scores show these predictors are uncorrelated.

### Cluster Analysis for Brain Correlates of Tremor Severity

Two clusters of significant spatial extent were observed in the occipital (extent∼2800 voxels, *p*_k_< 1×10^-8^) and parietal (extent ∼1600 voxels, *p*_k_< 1×10^-6^) areas (Figure 3a), where *R*_1_ values were negatively correlated with tremor scores in PD subjects. Scatter plots between the mean *R*_1_ values within the clusters and tremor scores showed a high negative correlation between these measures in both clusters (occipital cluster: Pearson’s *r* = -0.65, *p* = 8.7×10^-5^, parietal cluster: Pearson’s *r* = -0.63, *p* = 1.8×10^-4^) without any major outliers (Figure 3b and 3c). No significant clusters were observed for the correlation of *BP*_ND_ with tremor scores.

**Fig. 3.**
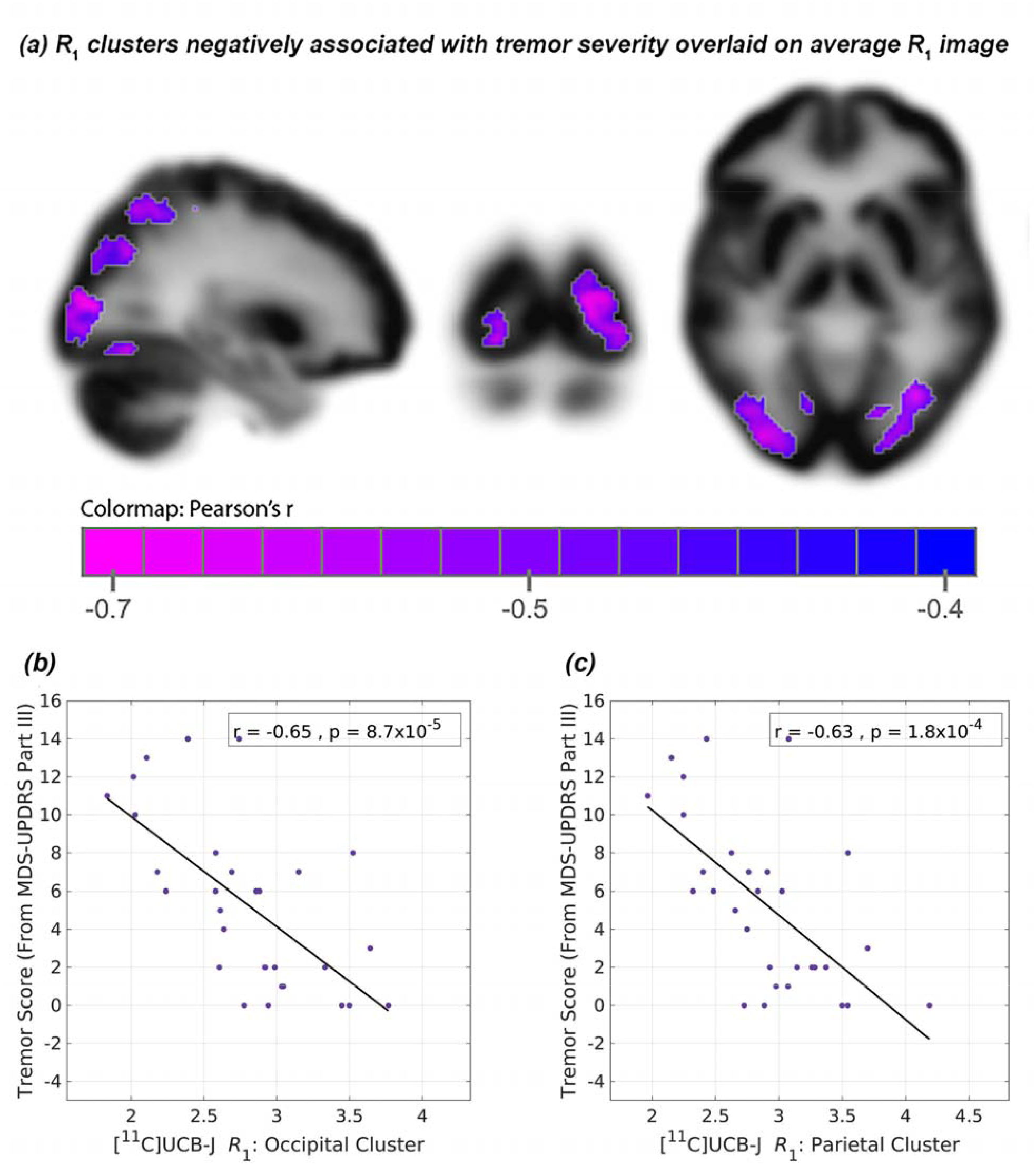
(a) Clusters of significant spatial extent observed within the occipital and parietal areas where *R*_1_ values were negatively correlated with tremor scores in PD subjects. The large spatial extents of these clusters were unlikely due to random chance (*p*_k_ < 1×10^-6^, spatial smoothness applied to the images accounted for in *p*_k_ calculation). (b, c) Significant negative correlations (across-PD subjects) between tremor scores and mean *R*_1_ values in the occipital and parietal clusters, respectively.

### Alterations of synaptic (BP_ND_) and perfusion (R_1_) correlations in PD

Compared to HC, synaptic correlations in PD were significantly lowered between the pallidum and many regions such as the brainstem (*d*_BP_= -0.47, *p*=0.03), red nucleus (*d*_BP_= -0.55, *p*=0.02), STN (*d*_BP_= -0.52, *p*=0.03) and SMA (*d*_BP_= -0.28, *p*=0.03). Synaptic association in PD was also lowered between the olfactory cortex and PCC (*d*_BP_= -0.29, *p*=0.01) but was higher between the raphe nucleus and thalamus (*d*_BP_=0.29, *p*=0.03). See Figure 4a and Supplementary Table S1.

**Fig. 4.**
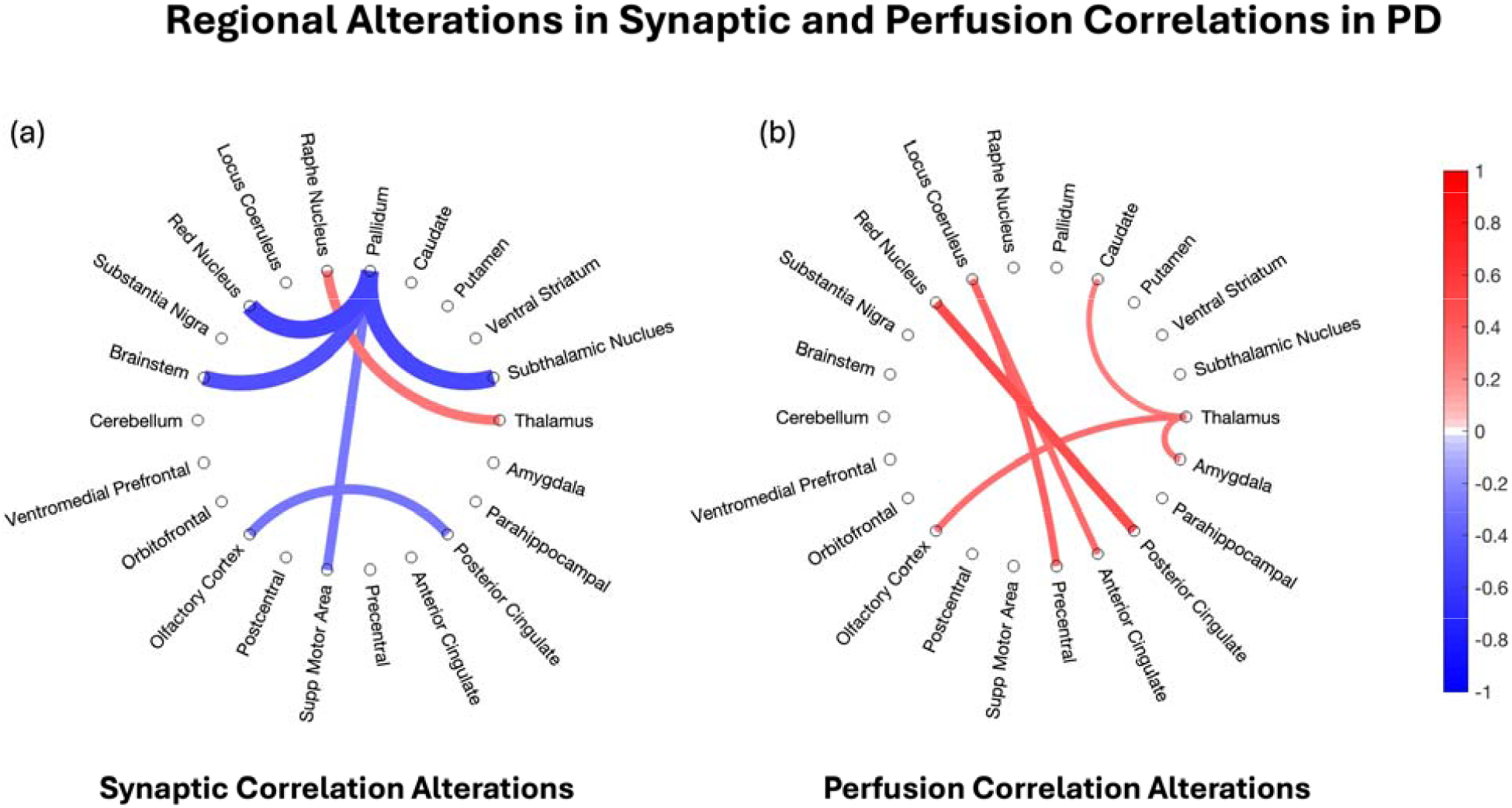
Group-level alterations in (a) synaptic (*BP*_ND_) and (b) perfusion (*R*_1_) correlations in PD compared to matched controls. Blue and red lines represent lower and higher change in correlations in PD, respectively, compared to controls. Line width and color intensities are proportional to the magnitude of the alteration in covariance, *d* = *r*_PD_ – *r*_HC_, where *r* is the Pearson’s correlation coefficient for either *BP*_ND_ or *R*_1_ across subjects for a given group and region-pair. *p* < 0.05 (non-parametric permutation test), uncorrected, considered significant for this exploratory analysis.

In contrast, most perfusion correlations were higher in PD between many ROI-pairs, for example between PCC and red nucleus (*d*_R1_=0.48, *p*=0.01). The thalamus was a hub of elevated perfusion associations with the caudate, amygdala and olfactory cortex (*d*_R1_ range=[0.26 0.32], *p* range=[0.006 0.01]). Similarly, the precentral cortex and ACC showed elevated associations with LC (*d*_R1_ range=[0.33 0.37], *p* range=[0.009 0.01]). See Figure 4b and Supplementary Table S2.

Scatter plots showing the correlation of the PET measure between the ROIs, separately in PD and controls, are available in Supplementary Figures S4 and S5 for each ROI-pair mentioned in the supplementary tables.

## Discussion

A major strength of [^11^C]UCB-J is its relatively high brain uptake due to which the *R*_1_ measure is a good surrogate for relative perfusion. Utilizing *R*_1_ images, we were able to derive the PDRP – an abnormal network associated with motor symptoms of PD - in our cohort. This pattern, first described by Eidelberg et al.[22], remains one of the most reproduced imaging biomarkers of PD and has been derived using several imaging modalities measuring blood flow[37-40] or glucose metabolism[23-25, 35-37]. Importantly, we further showed that by combining synaptic density losses in the substantia nigra as well as PDRP expression scores, a single [^11^C]UCB-J PET image can integrate complementary information and better explain motor severity variation in early-stage PD. We also found large clusters in occipital-parietal areas where relative blood blow was negatively correlated with tremor scores in PD. To gain further insight regarding the differences between synaptic and perfusion (*R*_1_) measures in PD, we performed an exploratory analysis characterizing alterations in regional synaptic and perfusion correlations. Our results showed that the two are indeed distinct; most synaptic alterations in PD led to lowered synaptic associations, but most significant perfusion changes involved elevated perfusion correlations between regions. Since these findings are novel, it is important to address them further regarding their interpretation, potential caveats and some suggestions for validation.

The PDRP identified in the present analysis was topographically similar to previous reports[23, 25, 37] and was not only expressed more in PD but subject scores were also correlated with MDS-UPDRS III scores. Yet, the percent variation explained by PC1 was only 7.4% which is considerably lower than 15-20% reported for PDRP derived from [^18^F]FDG PET[23, 25], though some previous reports for PDRP based on blood flow MRI measures have reported comparable values[39]. The correlation of PDRP expression with MDS-UPDRS III was also significant but modest. We ascribe these differences to (1) early and uniform disease staging of our PD cohort (Hoehn-Yahr 2) that diminishes the dynamic range of biological variability in our sample compared to previous studies, and (2) a higher-resolution PET camera with limited smoothing leading to finer features that could distribute the variance across a larger number of PCs. We studied whether additional smoothing or a different way of selecting voxels could increase overall %variation explained by the PDRP. These do produce a modest effect of 3-5%, likely by reducing total variance due to noise reduction, however, improving %variation by itself is not the goal of a PCA analysis. Rather, the aim is to find a reliable component that separates PD from controls and correlates better with the PD disease severity. In our main analysis, PC1 accomplishes this despite only explaining 7.4% of the variation. Generally, the degree of PDRP expression in PD patients and its correlation with MDS-UPDRS III depends on their disease stage and duration, with stronger expression in subjects with more severe PD[37, 41], and weaker expression in early and *de novo* stages of the disease[38, 42, 43].

It is also unsurprising that SSM-PCA analysis on *BP*_ND_ images did not find extensive patterns unique to PD. Our previous [^11^C]UCB-J PET study in this cohort[21] showed that synaptic losses are generally focal, with strongest effects in small mid-brain nuclei such as the substantia nigra, until later stages or higher disease durations. These focal differences are unlikely to stand out in a voxel-level PCA analysis across subjects, especially with smoothed images. However, we demonstrated that PDRP expression in PD and synaptic losses in the substantia nigra appear as complementary biological phenomena of motor severity worsening, such that their combination explains nearly 50% of the variation in motor impairment. We believe that this is impressive for an early clinical stage cohort and should be tested on *de novo* participants as well as in prodromal suspected cases to see if it has the potential to improve earlier diagnosis.

Next, cluster analyses showed that tremor severity in PD was negatively associated with relative perfusion in large areas within occipital and parietal lobes. This finding is surprising as current circuit models of tremor in PD implicate the subthalamic nucleus and the overactivity of cerebello-thamalo-cortical circuit[28] for the generation and amplification of tremor, respectively. One explanation could be due to the differences in physiological measurements enabled by [^11^C]UCB-J *R*_1_ and electrophysiology/fMRI on which the current circuit models of tremor are based. [^11^C]UCB-J *R*_1_ measures a static image of relative perfusion, while electrophysiology/fMRI data measure temporal fluctuations in BOLD signal. For a correlative analysis across subjects these two measures reflect on different physiological processes – temporal fluctuations in blood oxygenation for fMRI vs average brain perfusion at resting state for PET. Our results imply that amplification of tremor may have additional contributions from relative perfusion within the occipital and parietal lobes. We speculate that alterations in visual processing in the occipital lobe and proprioception-visual information integration in the parietal lobe could contribute to the development of tremor.

Finally, we performed an exploratory analysis to separately account for alterations in regional synaptic and perfusion (*R*_1_) correlations (or ‘regional associations’) in PD, at the group level. Our results suggest that synaptic associations between any two nodes can influence yet be distinct from functional (perfusion) associations. For example, an expected decrease in synaptic loss between two nodes (or regions) may be compensated by higher neuronal activity in that functional pathway, or through generation of other functional pathways between the two nodes. For synaptic correlations, the most striking feature was the reduced associations of the pallidum with the brainstem, red nucleus, STN and SMA, all of which are key regions of pathological motor circuitry in PD. Since the pallidum is an inhibitory output nucleus to the basal ganglia and thalamic regions[44], loss of synaptic associations between the pallidum and these regions may be partly responsible for the higher functional associativity between some thalamic nuclei and cortical regions in PD[45-47]. Taken together, our findings show a main correlation pattern of reduced synaptic associations and increased functional associations due to increased neurovascular coupling arising from neuronal disinhibition (likely explanation for the overactivity of thalamus) or compensatory overactivation. This analysis requires independent validation as corrections for multiple comparisons could not be made given the exploratory nature of findings.

## Conclusion

This study demonstrates the utility of [^11^C]UCB-J PET in PD beyond measuring synaptic loss. Dynamic imaging enabled computation of a measure of relative perfusion (*R*_1_) which was used to derive the PDRP-an abnormal metabolic network that was expressed more in PD participants and correlated with MDS-UPDRS Part III scores. Combining measures of focal synaptic losses in the substantia nigra and distributed metabolic pattern changes with PDRP dramatically improved the explanation of PD motor severity, showing how [^11^C]UCB-J provides two important biomarkers in a single-scan. Cluster analysis identified significantly large areas in the occipital-parietal regions where relative blood flow was significantly negatively correlated with tremor severity, a symptom that is generally unexplained by dopaminergic losses[27]. Finally, we reported remodeling of several unique alterations in regional synaptic and perfusion correlations in PD, at the group level, which may aid in understanding the large-scale associativity changes due to the disease processes, as well as potential compensation pathways.

## Acknowledgements

The work reported in the present manuscript was supported by funds from NIH (Matuskey, 1R01NS12481) and Abbvie, Inc. We acknowledge the use of Scan Analysis and Visualization Processor MATLAB toolbox, developed at Feinstein Institute for Medical Research by Dr. David Eidelberg and Dr. Phoebe Spetsieris.

## Data Availability

Raw data on which this work is based is available from the principal investigator (Dr. David Matuskey) upon request.

## References

1. Dorsey ER, Sherer T, Okun MS, Bloem BR. The Emerging Evidence of the Parkinson Pandemic. J Parkinsons Dis. 2018;8:S3–S8. doi:10.3233/JPD-181474.

2. Spillantini MG, Schmidt ML, Lee VM, Trojanowski JQ, Jakes R, Goedert M. Alpha-synuclein in Lewy bodies. Nature. 1997;388:839–40. doi:10.1038/42166.

3. Maroteaux L, Campanelli JT, Scheller RH. Synuclein: a neuron-specific protein localized to the nucleus and presynaptic nerve terminal. J Neurosci. 1988;8:2804–15. doi:10.1523/JNEUROSCI.08-08-02804.1988.

4. Burré JS, M.; Südhof, T. C. α-Synuclein assembles into higher-order multimers upon membrane binding to promote SNARE complex formation. Proc Natl Acad Sci USA. 2014;111(40):E4274–E83.

5. Gao V, Briano JA, Komer LE, Burre J. Functional and Pathological Effects of alpha-Synuclein on Synaptic SNARE Complexes. J Mol Biol. 2023;435:167714. doi:10.1016/j.jmb.2022.167714.

6. Vargas KJS, N.; Davis T.; Fernandez-Busnadiego, R.; Taguchi, Y.V.; Laugks, U.; Lucic, V.; Chandra, S. S. Synucleins have multiple effects on presynaptic architecture. Cell Rep. 2017;18:161–73.

7. Nabulsi NB, Mercier J, Holden D, Carre S, Najafzadeh S, Vandergeten MC, et al. Synthesis and Preclinical Evaluation of 11C-UCB-J as a PET Tracer for Imaging the Synaptic Vesicle Glycoprotein 2A in the Brain. J Nucl Med. 2016;57:777–84. doi:10.2967/jnumed.115.168179.

8. Mercier J, Archen L, Bollu V, Carre S, Evrard Y, Jnoff E, et al. Discovery of heterocyclic nonacetamide synaptic vesicle protein 2A (SV2A) ligands with single-digit nanomolar potency: opening avenues towards the first SV2A positron emission tomography (PET) ligands. ChemMedChem. 2014;9:693–8. doi:10.1002/cmdc.201300482.

9. Li S, Cai Z, Wu X, Holden D, Pracitto R, Kapinos M, et al. Synthesis and in Vivo Evaluation of a Novel PET Radiotracer for Imaging of Synaptic Vesicle Glycoprotein 2A (SV2A) in Nonhuman Primates. ACS Chem Neurosci. 2019;10:1544–54. doi:10.1021/acschemneuro.8b00526.

10. Cai Z, Li S, Zhang W, Pracitto R, Wu X, Baum E, et al. Synthesis and Preclinical Evaluation of an (18)F-Labeled Synaptic Vesicle Glycoprotein 2A PET Imaging Probe: [(18)F]SynVesT-2. ACS Chem Neurosci. 2020;11:592–603.doi:10.1021/acschemneuro.9b00618.

11. Finnema SJ, Nabulsi NB, Eid T, Detyniecki K, Lin SF, Chen MK, et al. Imaging synaptic density in the living human brain. Sci Transl Med. 2016;8:348ra96. doi:10.1126/scitranslmed.aaf6667.

12. Finnema SJ, Nabulsi NB, Mercier J, Lin SF, Chen MK, Matuskey D, et al. Kinetic evaluation and test-retest reproducibility of [(11)C]UCB-J, a novel radioligand for positron emission tomography imaging of synaptic vesicle glycoprotein 2A in humans. J Cereb Blood Flow Metab. 2018;38:2041–52. doi:10.1177/0271678×17724947.

13. Naganawa M, Li S, Nabulsi N, Henry S, Zheng MQ, Pracitto R, et al. First-in-Human Evaluation of (18)F-SynVesT-1, a Radioligand for PET Imaging of Synaptic Vesicle Glycoprotein 2A. J Nucl Med. 2021;62:561–7. doi:10.2967/jnumed.120.249144.

14. Li S, Naganawa M, Pracitto R, Najafzadeh S, Holden D, Henry S, et al. Assessment of test-retest reproducibility of [(18)F]SynVesT-1, a novel radiotracer for PET imaging of synaptic vesicle glycoprotein 2A. Eur J Nucl Med Mol Imaging. 2021;48:1327–38. doi:10.1007/s00259-020-05149-3.

15. Matuskey D, Tinaz S, Wilcox KC, Naganawa M, Toyonaga T, Dias M, et al. Synaptic Changes in Parkinson Disease Assessed with in vivo Imaging. Ann Neurol. 2020;87:329–38. doi:10.1002/ana.25682.

16. Delva A, Van Weehaeghe D, Koole M, Van Laere K, Vandenberghe W. Loss of Presynaptic Terminal Integrity in the Substantia Nigra in Early Parkinson’s Disease. Mov Disord. 2020;35:1977–86. doi:10.1002/mds.28216.

17. Wilson H, Pagano G, de Natale ER, Mansur A, Caminiti SP, Polychronis S, et al. Mitochondrial Complex 1, Sigma 1, and Synaptic Vesicle 2A in Early Drug-Naive Parkinson’s Disease. Mov Disord. 2020;35:1416–27. doi:10.1002/mds.28064.

18. Delva A, Van Laere K, Vandenberghe W. Longitudinal Positron Emission Tomography Imaging of Presynaptic Terminals in Early Parkinson’s Disease. Mov Disord. 2022;37:1883–92. doi:10.1002/mds.29148.

19. Andersen KB, Hansen AK, Damholdt MF, Horsager J, Skjaerbaek C, Gottrup H, et al. Reduced Synaptic Density in Patients with Lewy Body Dementia: An [(11) C]UCB-J PET Imaging Study. Mov Disord. 2021;36:2057–65. doi:10.1002/mds.28617.

20. Andersen KB, Hansen AK, Schacht AC, Horsager J, Gottrup H, Klit H, et al. Synaptic Density and Glucose Consumption in Patients with Lewy Body Diseases: An [(11) C]UCB-J and [(18) F]FDG PET Study. Mov Disord. 2023;38:796–805. doi:10.1002/mds.29375.

21. Holmes SE, Honhar P, Tinaz S, Naganawa M, Hilmer AT, Gallezot JD, et al. Synaptic loss and its association with symptom severity in Parkinson’s disease. NPJ Parkinsons Dis. 2024;10:42. doi:10.1038/s41531-024-00655-9.

22. Eidelberg D, Moeller JR, Dhawan V, Spetsieris P, Takikawa S, Ishikawa T, et al. The metabolic topography of parkinsonism. J Cereb Blood Flow Metab. 1994;14:783–801. doi:10.1038/jcbfm.1994.99.

23. Spetsieris PG, Eidelberg D. Scaled subprofile modeling of resting state imaging data in Parkinson’s disease: methodological issues. Neuroimage. 2011;54:2899–914. doi:10.1016/j.neuroimage.2010.10.025.

24. Teune LK, Renken RJ, de Jong BM, Willemsen AT, van Osch MJ, Roerdink JB, et al. Parkinson’s disease-related perfusion and glucose metabolic brain patterns identified with PCASL-MRI and FDG-PET imaging. Neuroimage Clin. 2014;5:240–4. doi:10.1016/j.nicl.2014.06.007.

25. Meles SK, Renken RJ, Pagani M, Teune LK, Arnaldi D, Morbelli S, et al. Abnormal pattern of brain glucose metabolism in Parkinson’s disease: replication in three European cohorts. Eur J Nucl Med Mol Imaging. 2020;47:437–50. doi:10.1007/s00259-019-04570-7.

26. Moeller JR, Strother SC. A regional covariance approach to the analysis of functional patterns in positron emission tomographic data. J Cereb Blood Flow Metab. 1991;11:A121–35. doi:10.1038/jcbfm.1991.47.

27. Kerstens VS, Fazio P, Sundgren M, Halldin C, Svenningsson P, Varrone A. [(18)F]FE-PE2I DAT correlates with Parkinson’s disease duration, stage, and rigidity/bradykinesia scores: a PET radioligand validation study. EJNMMI Res. 2023;13:29. doi:10.1186/s13550-023-00974-7.

28. Buijink AWG, van Rootselaar AF, Helmich RC. Connecting tremors - a circuits perspective. Curr Opin Neurol. 2022;35:518–24. doi:10.1097/WCO.0000000000001071.

29. Postuma RB, Berg D, Stern M, Poewe W, Olanow CW, Oertel W, et al. MDS clinical diagnostic criteria for Parkinson’s disease. Mov Disord. 2015;30:1591–601. doi:10.1002/mds.26424.

30. Goetz CG, Tilley BC, Shaftman SR, Stebbins GT, Fahn S, Martinez-Martin P, et al. Movement Disorder Society-sponsored revision of the Unified Parkinson’s Disease Rating Scale (MDS-UPDRS): scale presentation and clinimetric testing results. Mov Disord. 2008;23:2129–70. doi:10.1002/mds.22340.

31. Stebbins GT, Goetz CG, Burn DJ, Jankovic J, Khoo TK, Tilley BC. How to identify tremor dominant and postural instability/gait difficulty groups with the movement disorder society unified Parkinson’s disease rating scale: comparison with the unified Parkinson’s disease rating scale. Mov Disord. 2013;28:668–70. doi:10.1002/mds.25383.

32. Jin X, Mulnix T, Gallezot JD, Carson RE. Evaluation of motion correction methods in human brain PET imaging--a simulation study based on human motion data. Med Phys. 2013;40:102503. doi:10.1118/1.4819820.

33. Wu Y, Carson RE. Noise reduction in the simplified reference tissue model for neuroreceptor functional imaging. J Cereb Blood Flow Metab. 2002;22:1440–52. doi:10.1097/01.WCB.0000033967.83623.34.

34. Rossano S, Toyonaga T, Finnema SJ, Naganawa M, Lu Y, Nabulsi N, et al. Assessment of a white matter reference region for (11)C-UCB-J PET quantification. J Cereb Blood Flow Metab. 2020;40:1890–901. doi:10.1177/0271678×19879230.

35. Spetsieris P, Ma Y, Peng S, Ko JH, Dhawan V, Tang CC, et al. Identification of disease-related spatial covariance patterns using neuroimaging data. J Vis Exp. 2013. doi:10.3791/50319.

36. Ko JH, Katako A, Aljuaid M, Goertzen AL, Borys A, Hobson DE, et al. Distinct brain metabolic patterns separately associated with cognition, motor function, and aging in Parkinson’s disease dementia. Neurobiol Aging. 2017;60:81–91. doi:10.1016/j.neurobiolaging.2017.08.020.

37. Ma Y, Tang C, Spetsieris PG, Dhawan V, Eidelberg D. Abnormal metabolic network activity in Parkinson’s disease: test-retest reproducibility. J Cereb Blood Flow Metab. 2007;27:597–605. doi:10.1038/sj.jcbfm.9600358.

38. Peng S, Tang C, Schindlbeck K, Rydzinski Y, Dhawan V, Spetsieris PG, et al. Dynamic (18)F-FPCIT PET: Quantification of Parkinson’s disease metabolic networks and nigrostriatal dopaminergic dysfunction in a single imaging session. J Nucl Med. 2021;62:1775–82. doi:10.2967/jnumed.120.257345.

39. Rane S, Koh N, Oakley J, Caso C, Zabetian CP, Cholerton B, et al. Arterial spin labeling detects perfusion patterns related to motor symptoms in Parkinson’s disease. Parkinsonism Relat Disord. 2020;76:21–8. doi:10.1016/j.parkreldis.2020.05.014.

40. Vo A, Sako W, Fujita K, Peng S, Mattis PJ, Skidmore FM, et al. Parkinson’s disease-related network topographies characterized with resting state functional MRI. Hum Brain Mapp. 2017;38:617–30. doi:10.1002/hbm.23260.

41. Huang C, Tang C, Feigin A, Lesser M, Ma Y, Pourfar M, et al. Changes in network activity with the progression of Parkinson’s disease. Brain. 2007;130:1834–46. doi:10.1093/brain/awm086.

42. Matthews DC, Lerman H, Lukic A, Andrews RD, Mirelman A, Wernick MN, et al. FDG PET Parkinson’s disease-related pattern as a biomarker for clinical trials in early stage disease. Neuroimage Clin. 2018;20:572–9. doi:10.1016/j.nicl.2018.08.006.

43. Schindlbeck KA, Lucas-Jimenez O, Tang CC, Morbelli S, Arnaldi D, Pardini M, et al. Metabolic Network Abnormalities in Drug-Naive Parkinson’s Disease. Mov Disord. 2020;35:587–94. doi:10.1002/mds.27960.

44. Dong J, Hawes S, Wu J, Le W, Cai H. Connectivity and Functionality of the Globus Pallidus Externa Under Normal Conditions and Parkinson’s Disease. Front Neural Circuits. 2021;15:645287. doi:10.3389/fncir.2021.645287.

45. Owens-Walton C, Jakabek D, Power BD, Walterfang M, Velakoulis D, van Westen D, et al. Increased functional connectivity of thalamic subdivisions in patients with Parkinson’s disease. PLoS One. 2019;14:e0222002. doi:10.1371/journal.pone.0222002.

46. Agosta F, Caso F, Stankovic I, Inuggi A, Petrovic I, Svetel M, et al. Cortico-striatal-thalamic network functional connectivity in hemiparkinsonism. Neurobiol Aging. 2014;35:2592–602. doi:10.1016/j.neurobiolaging.2014.05.032.

47. Zeng Q, Guan X, Guo T, Law Yan Lun JCF, Zhou C, Luo X, et al. The Ventral Intermediate Nucleus Differently Modulates Subtype-Related Networks in Parkinson’s Disease. Front Neurosci. 2019;13:202. doi:10.3389/fnins.2019.00202.

