## Supplementary for "Alterations in relative perfusion and synaptic patterns in Parkinson’s disease using dynamic SV2A PET"

*Supplemental Information*

### *1. Optimization of lower (cutoff) threshold for PDRP - network analysis with [$^{11}$C]UCB-J PET*

According to the methodological description of SSM-PCA for [$^{18}$F]FDG PET data by Spetsieris et al.[1], a lower cutoff threshold (in units of % of maximum voxel value) was recommended to create masks for individual scans, that can be multiplicatively combined to define a group-level common mask which is then used to remove out of brain-voxels and voxels with low activity (low signal-to-background) before applying the SSM-PCA algorithm. Their study recommended a value of 35% of the maximum voxel value as the optimum lower threshold for [$^{18}$F]FDG PET images based on highest separation of controls from Parkinson's disease (PD) subjects from the computed PDRP for this threshold[1]. However, since [$^{11}$C]UCB-J PET is a new modality for SSM-PCA, we optimized this threshold mask again following the methods of Spetsieris et al[1].

We investigated six different values for the lower (cutoff) threshold (in units of % of maximum voxel value) – 25, 30, 35, 40, 45 and 50 – resulting in six different group-level masks of acceptable voxels, that were each analyzed separately with SSM-PCA analysis to derive PDRP in each case. It should be noted that a smaller value for lower threshold would include more voxels and hence result in a larger mask. The optimum threshold was defined on the basis of the significance level ($p$-value) of PDRP expression scores in discriminating PD subjects from controls. Note that $R_1$ images are generally noisier than early standardized uptake value ratio ($SUVR$) images, and maximum voxel $R_1$ value in the image can be quite high due to noise in the model– therefore, this analysis was performed using the early $SUVR$ (0-10 min) instead.

In each case, the first principal component from SSM-PCA analysis of early $SUVR$ images (0-10 min) was identified as the PDRP through visualization of its loading weights. For almost every case (except threshold=50%), the subject expression scores of the network was significantly different between controls and PD subjects (higher in PD subjects, Figure S1). The lowest $p$-value for the group difference (best group separation) was achieved for the mask corresponding to the lower threshold value equal to 30% of maximum voxel value. The optimized mask for values greater than this threshold, shown in Figure S2, is visually similar to a mask for gray-matter voxels. This optimized mask was also used in the main analysis with $R_1$ images to remove low activity voxels as reported in the manuscript.

The optimized PDRP based on $SUVR$ [0-10 min] ($PDRP_{SUVR}$) is shown in Figure S3. Not only does it visually compare well against the PDRP computed from $R_1$ (Figure 1 in manuscript), but $z$-scores of $PDRP_{SUVR}$ expression also discriminated the control and PD groups ($z$-scores for controls: -0.47±0.86; for PD: 0.45±0.93, $p = 3.7x10^{-5}$, Cohen's $d = 1.0$) to similar extent as the PDRP derived from $R_1$ ($z$-scores for controls: -0.52±0.73; for PD: 0.52±0.97, $p = 1.7x10^{-5}$, Cohen's $d = 1.2$). Additionally, $PDRP_{SUVR}$ expression scores in individuals with PD were also significantly correlated with MDS-UPDRS Part III (Pearson's $r$ = 0.54, $p = 0.002$ vs $r = 0.57$, $p = 0.0009$ for PDRP based on $R_1$). Finally, the $z$-scores for $PDRP_{SUVR}$ and PDRP-$R_1$ expression were strongly correlated in PD (Pearson's $r = 0.99$, $p < 1x10^{-16}$) and controls (Pearson's $r = 0.97$, $p < 1x10^{-16}$), demonstrating that these measures are practically interchangeable.

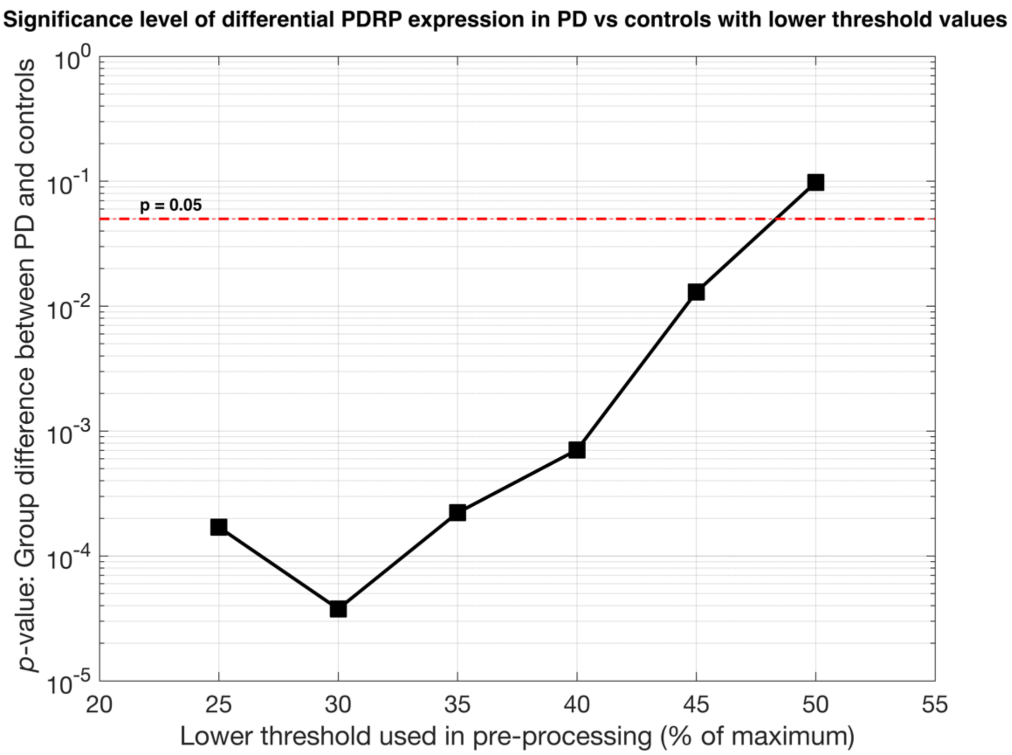


Figure S1: *p* values for the difference in PDRP expression (PDRP from *SUVR* 0-10 min) between PD and controls as a function of different lower (cutoff) thresholds used in data pre-processing. A lower threshold corresponding 30% of maximum value was found to be the optimum choice for selecting voxels for SSM-PCA analysis to ensure best separation of PD from controls.

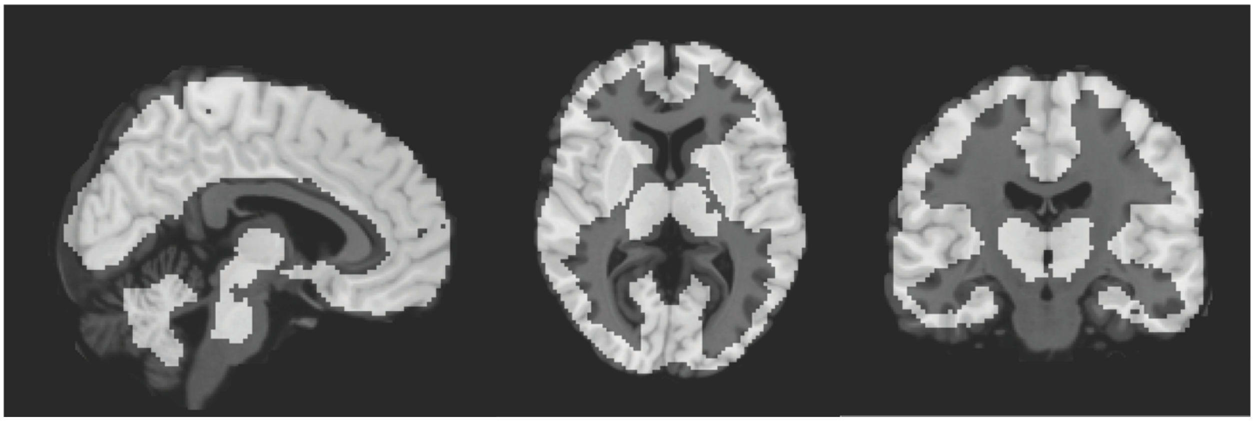

Figure S2: Optimal brain mask for [$^{11}$C]UCB-J SSM-PCA analysis (overlay, white) created from PET data by only including voxels with *SUVR* 0-10 min greater than 30% of the group-level maximum voxel value for *SUVR* 0-10 min value across all subjects in the analysis. The mask, overlaid on a 1mm T1 MR image for comparison, includes most gray-matter voxels.

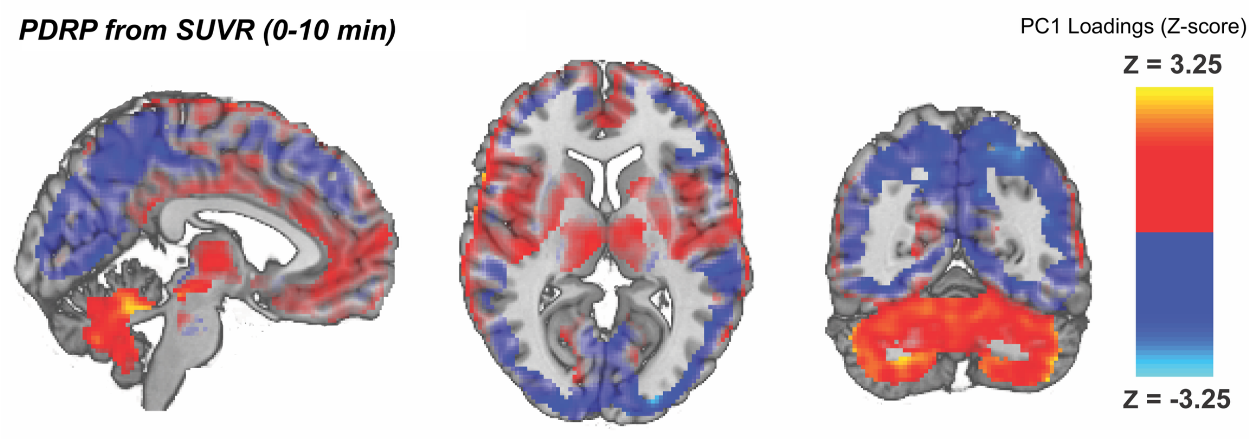


Figure S3: PDRP derived from *SUVR* 0-10 images with the optimal lower threshold (30% of group-level maximum value for *SUVR*) during pre-processing step, was very similar to those derived from $R_1$ images (Figure 1a in manuscript). The subject-level *z*-scores for expression of this network were strongly correlated with the corresponding values from PDRP-$R_1$ (Pearson's r > 0.97) and showed a comparable level of performance in differentiating PD from controls, and in their correlation with MDS-UPDRS Part III scores in the PD cohort (see Supplemental Material, Section 1, above).

### *More information on synaptic ($BP_{ND}$) and functional ($R_1$) covariance changes in PD*

**Table S1: List of significant alterations in synaptic ($BP_{ND}$) covariance at group-level in PD**

| S.No. | Region-Pair | $r_{BP,HC}$ | $r_{BP,PD}$ | $d_{BP} = r_{BP,PD} - r_{BP,HC}$ | p (permutation test) |
|---|---|---|---|---|---|
| 1 | Pallidum/Red Nucleus | 0.69 | 0.14 | -0.55 | 0.02 |
| 2 | Pallidum/Subthalamic Nucleus | 0.65 | 0.13 | -0.52 | 0.03 |
| 3 | Pallidum/Brainstem | 0.53 | 0.06 | -0.47 | 0.03 |
| 4 | Olfactory Cortex/Posterior Cingulate | 0.71 | 0.42 | -0.29 | 0.01 |
| 5 | Pallidum/Supplementary Motor Area | 0.79 | 0.51 | -0.28 | 0.03 |
| 6 | Thalamus/Raphe Nucleus | 0.52 | 0.81 | 0.29 | 0.03 |

***Table S2: List of significant alterations in functional ($R_1$)* connectivity at group-level in PD**

| S.No. | Region-Pair | $r_{R1,HC}$ | $r_{R1,PD}$ | $d_{R1} = r_{R1,PD} - r_{R1,HC}$ | *p* (permutation test) |
|---|---|---|---|---|---|
| 1 | Thalamus/Caudate | 0.65 | 0.91 | 0.26 | 0.006 |
| 2 | Thalamus/Amygdala | 0.53 | 0.82 | 0.29 | 0.009 |
| 3 | Thalamus/Olfactory Cortex | 0.54 | 0.86 | 0.32 | 0.01 |
| 4 | Locus Coeruleus/Anterior Cingulate | 0.22 | 0.55 | 0.33 | 0.009 |
| 5 | Locus Coeruleus/Precentral | 0.19 | 0.56 | 0.37 | 0.01 |
| 6 | Red Nucleus/Posterior Cingulate | 0.12 | 0.60 | 0.48 | 0.02 |

**Figure S4 (A-F): List of Pearson's correlation plots (separately for PD and HC) for all region-pairs with significant alterations in synaptic covariance**

**A:**

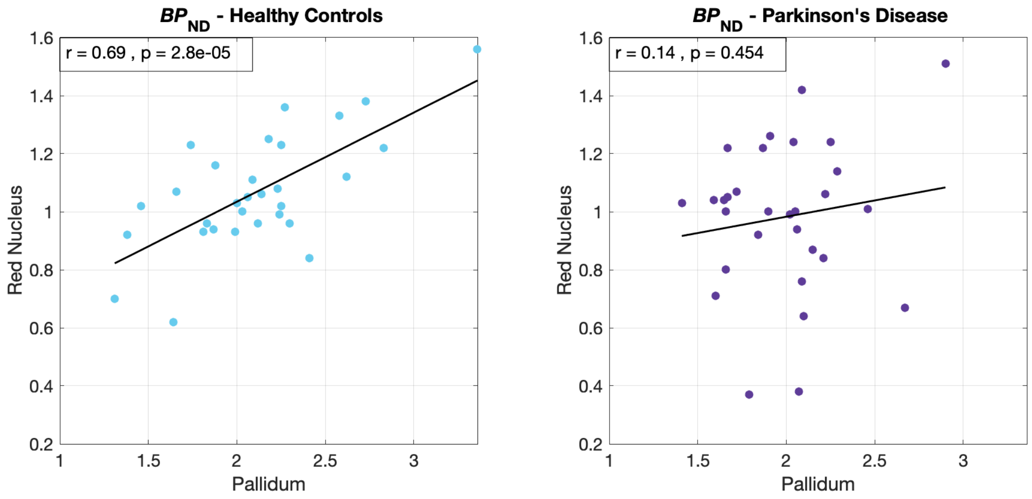


**B:**

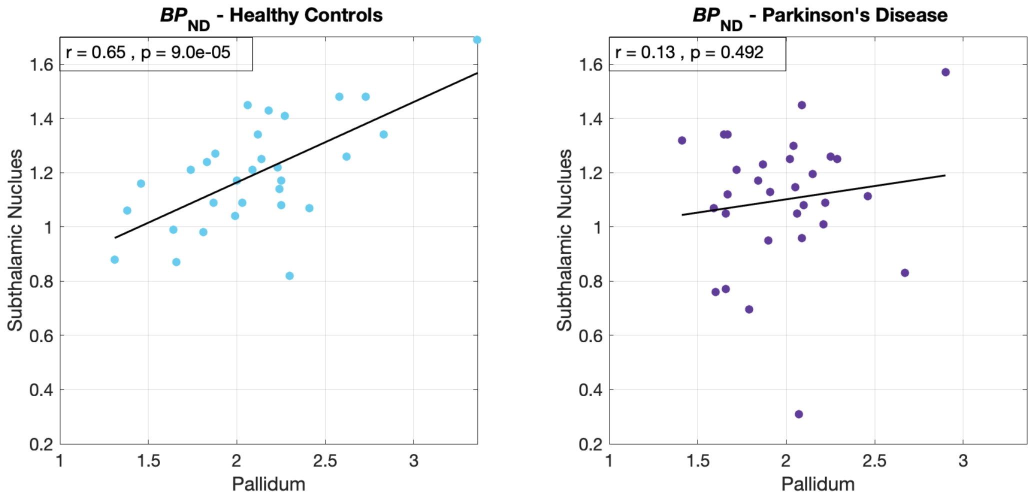

**C:**

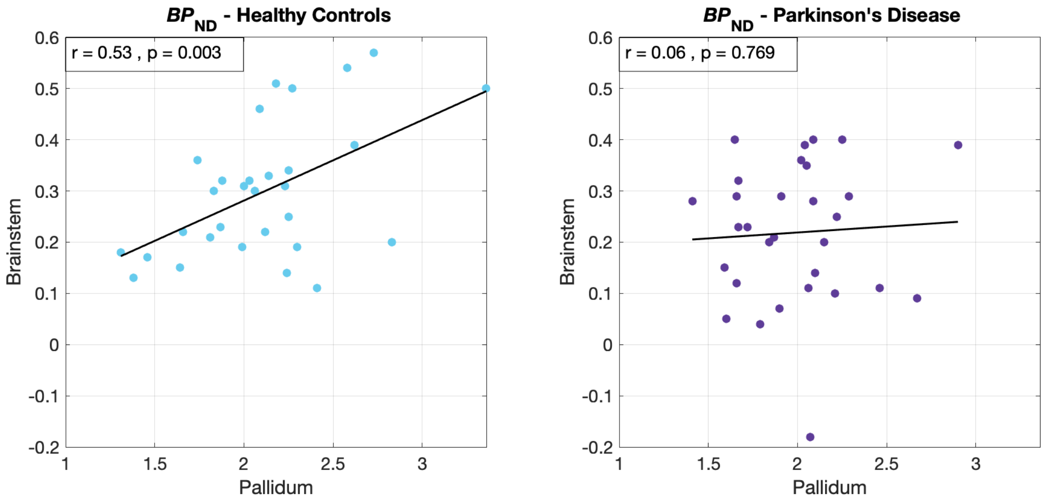
BPND - Healthy Controls
r = 0.53 , p = 0.003
Brainstem
Pallidum
BPND - Parkinson's Disease
r = 0.06 , p = 0.769
Brainstem
Pallidum

**D:**

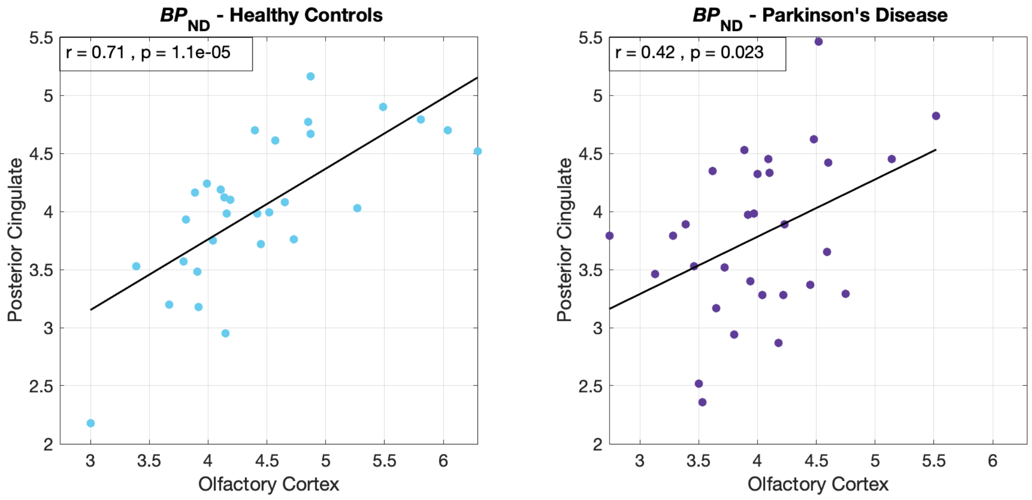
BPND - Healthy Controls
r = 0.71 , p = 1.1e-05
Posterior Cingulate
Olfactory Cortex
BPND - Parkinson's Disease
r = 0.42 , p = 0.023
Posterior Cingulate
Olfactory Cortex

**E:**

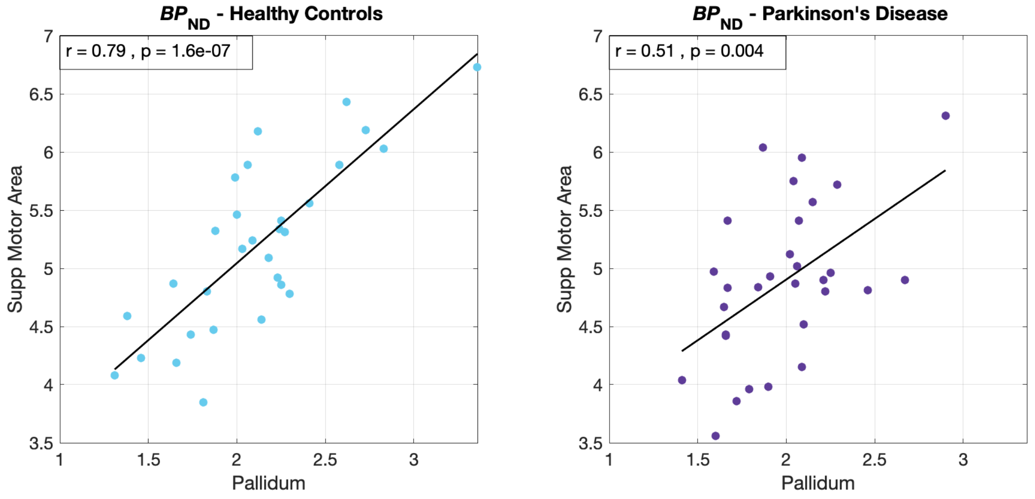

$BP_{ND}$ - Healthy Controls
r = 0.79 , p = 1.6e-07
Supp Motor Area
Pallidum
$BP_{ND}$ - Parkinson's Disease
r = 0.51 , p = 0.004
Supp Motor Area
Pallidum


**F:**

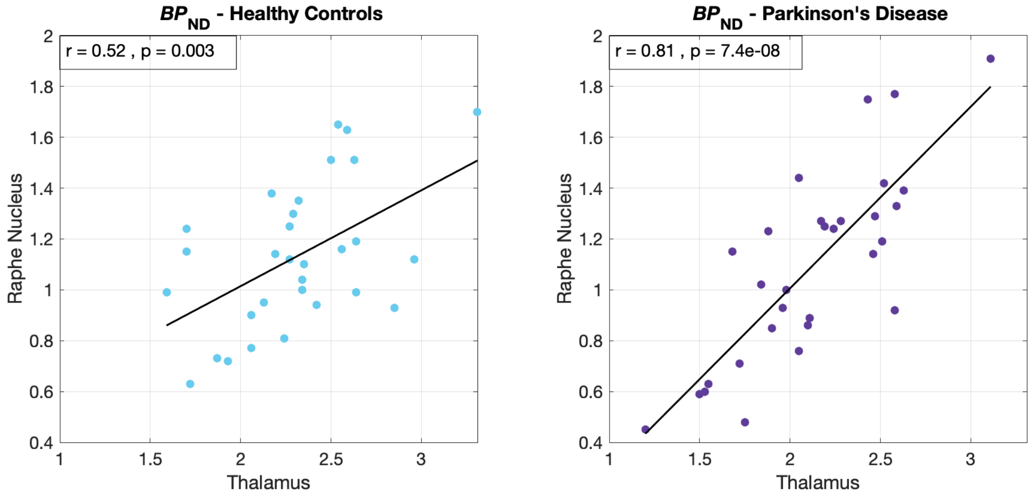

$BP_{ND}$ - Healthy Controls
r = 0.52 , p = 0.003
Raphe Nucleus
Thalamus
$BP_{ND}$ - Parkinson's Disease
r = 0.81 , p = 7.4e-08
Raphe Nucleus
Thalamus

**Figure S5 (A-F): List of Pearson's correlation plots (separately for PD and HC) for all region-pairs with significant alterations in regional functional covariance**

**A:**

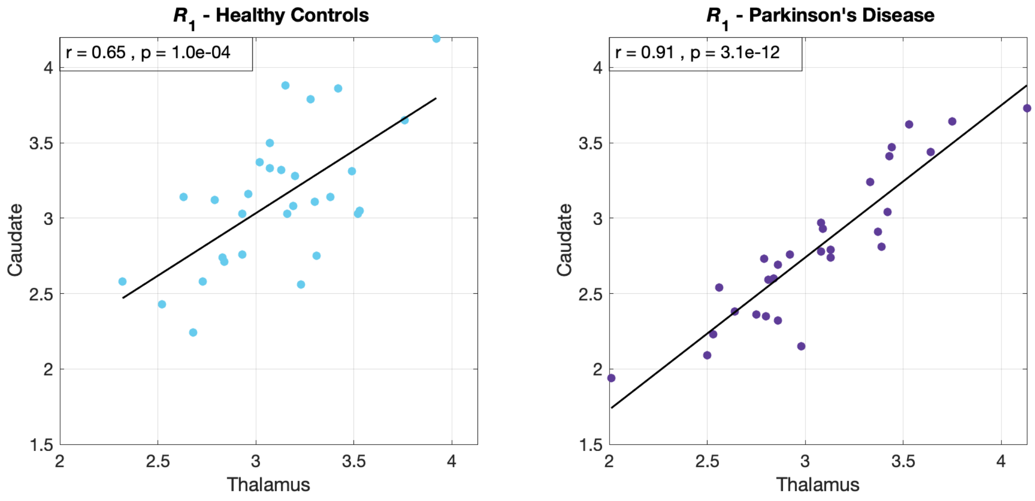


**B:**

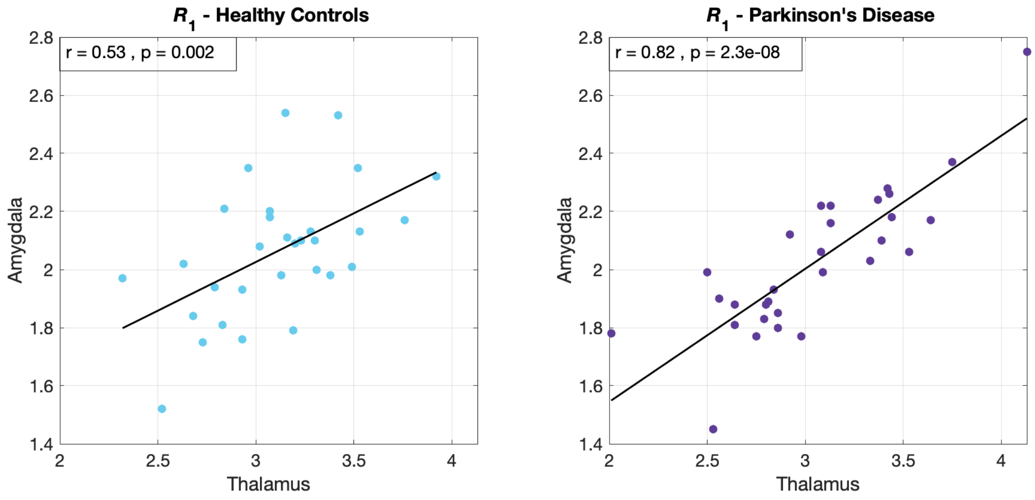

**C:**

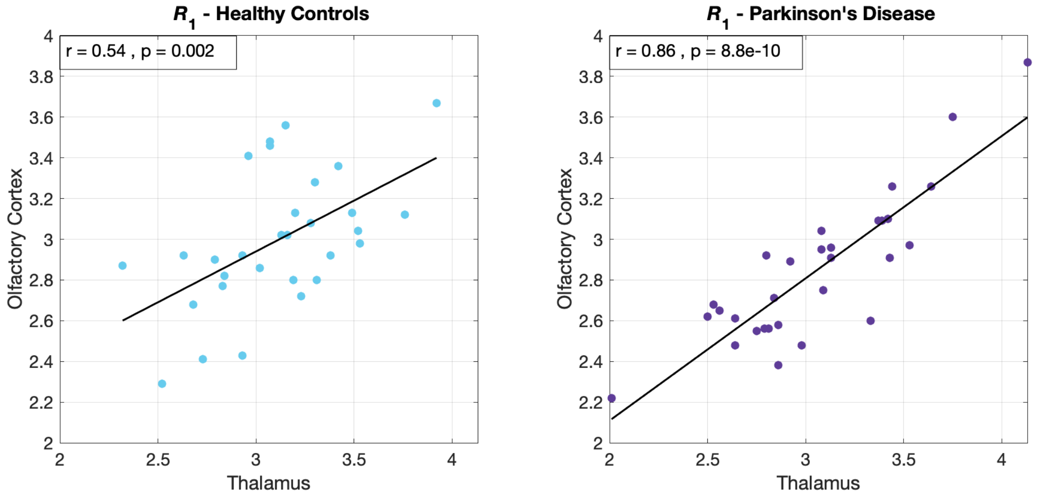

$R_1$ - Healthy Controls
r = 0.54 , p = 0.002
Olfactory Cortex
Thalamus
$R_1$ - Parkinson's Disease
r = 0.86 , p = 8.8e-10
Olfactory Cortex
Thalamus


**D:**

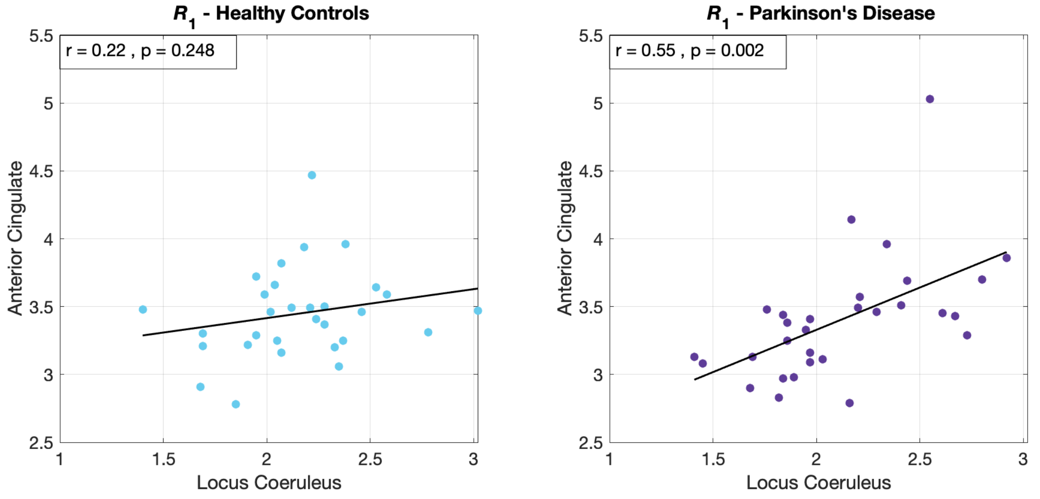

$R_1$ - Healthy Controls
r = 0.22 , p = 0.248
Anterior Cingulate
Locus Coeruleus
$R_1$ - Parkinson's Disease
r = 0.55 , p = 0.002
Anterior Cingulate
Locus Coeruleus

**E:**

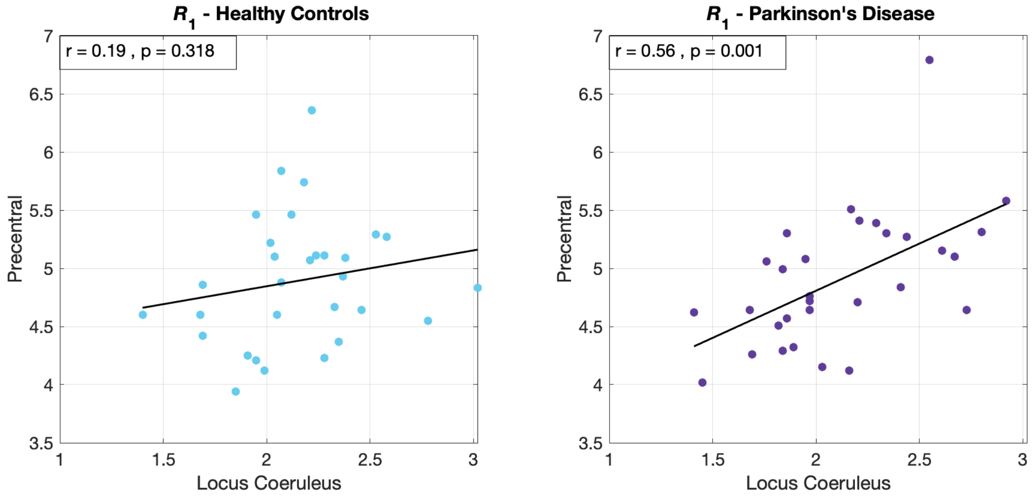


**F:**

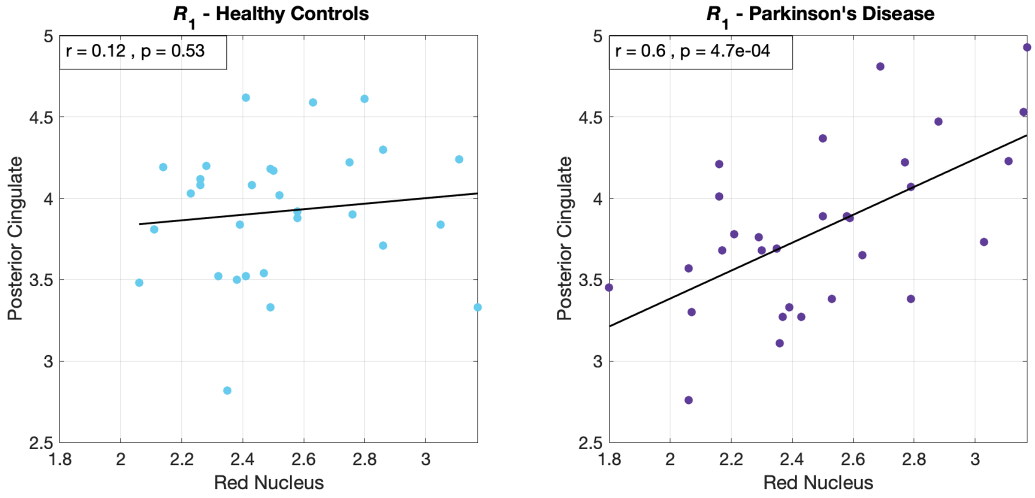


#### 3. *Tremor correlates using regional $R_1$ values for Parietal and Occipital lobes*

The cluster analyses found significant clusters within the occipital-parietal areas that were correlated with tremor severity in PD (Pearson's $r \sim -0.65$, $p < 0.001$). Therefore, we additionally investigated whether this association could be seen even from regional $R_1$ values. Regions of interest (ROIs) for the occipital and parietal lobes were defined using the Automated Anatomical Labelling atlas[2], and were then transformed to PET images of individuals using previously published methods[3]. Mean values of $R_1$ in the occipital and parietal ROIs were calculated and correlated with tremor scores.

Tremor scores showed a significant, but lowered, correlation with occipital ROI $R_1$ values (Pearson's $r = -0.38$, $p = 0.04$, Figure S4a) and non-significant negative correlation with parietal ROI $R_1$ values (Pearson's $r = -0.25$, $p = 0.19$, Figure S4b).

Therefore, voxel-wise $R_1$ cluster analysis provides higher sensitivity than regional analysis by detecting clusters that are most relevant to the tremor symptom within the relatively large occipital-parietal area.

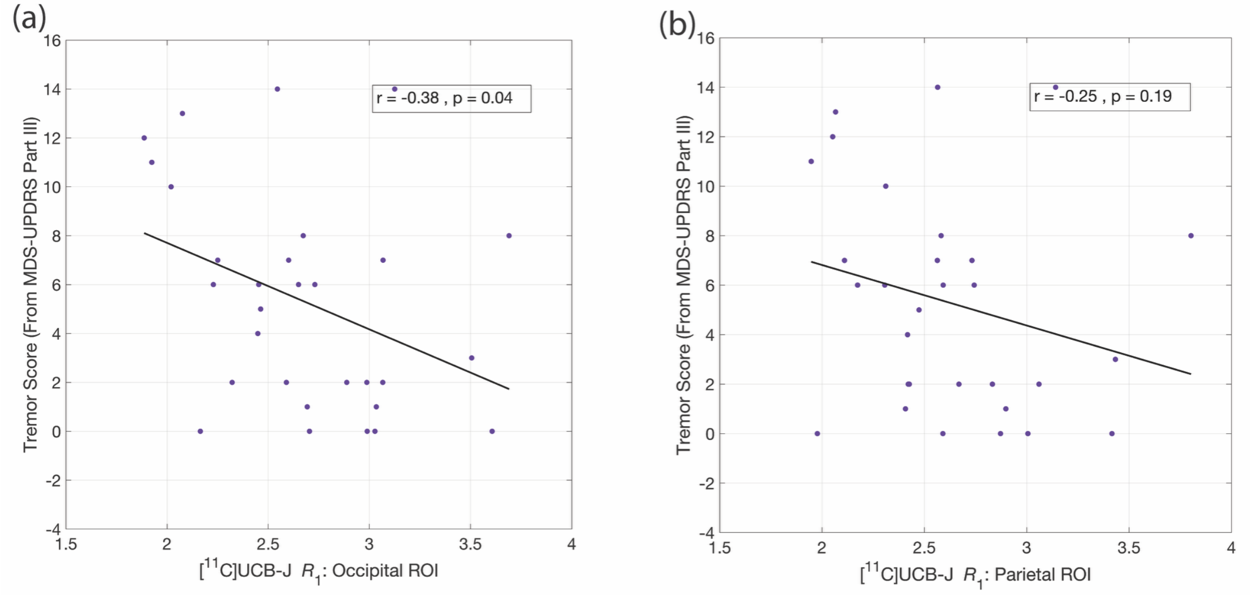


Figure S6: Correlation analysis between tremor severity with regional $R_1$ values in the occipital and parietal lobes showed lower correlation coefficients and statistical significance compared to the cluster analysis.

### *References*


1. Spetsieris PG, Eidelberg D. Scaled subprofile modeling of resting state imaging data in Parkinson's disease: methodological issues. *Neuroimage*. Feb 14 2011;54(4):2899-914. doi:10.1016/j.neuroimage.2010.10.025
2. Tzourio-Mazoyer N, Landeau B, Papathanassiou D, et al. Automated anatomical labeling of activations in SPM using a macroscopic anatomical parcellation of the MNI MRI single-subject brain. *Neuroimage*. Jan 2002;15(1):273-89. doi:10.1006/nimg.2001.0978
3. Matuskey D, Tinaz S, Wilcox KC, et al. Synaptic Changes in Parkinson Disease Assessed with in vivo Imaging. *Ann Neurol*. Mar 2020;87(3):329-338. doi:10.1002/ana.25682
